# Probing Hidden States for Calibrated, Alignment-Resistant Predictions in LLMs

**DOI:** 10.1101/2025.09.17.25336018

**Authors:** Jacob Berkowitz, Sophia Kivelson, Apoorva Srinivasan, Undina Gisladottir, Kevin K. Tsang, Jose Miguel Acitores Cortina, Aditi Kuchi, Jake Patock, Ryan Czarny, Nicholas P. Tatonetti

**Affiliations:** Department of Computational Biomedicine, Cedars-Sinai Medical Center, Los Angeles, 90069, CA, USA; Cedars-Sinai Cancer, Cedars-Sinai Medical Center, Los Angeles, 90069, CA, USA; Department of Biomedical Informatics, Columbia University Irving Medical Center, New York, 10032, NY, USA

**Keywords:** Large language models, Probing hidden states, Model calibration, Alignment resistance, Biomedical natural language processing

## Abstract

Scientific applications of large language models (LLMs) demand reliable, well-calibrated predictions, but standard generative approaches often fail to fully access relevant knowledge contained in their internal representations. As a result, models appear less capable than they are, with useful information remaining latent. We present PING (Probing INternal states of Generative models), an open-source framework that trains lightweight probes on frozen, HuggingFace-compatible transformers to deliver structured predictions with minimal compute overhead. Across diverse models and benchmarks including MMLU for broad coverage and MedMCQA for clinical focus, PING matches or exceeds generative accuracy while reducing Expected Calibration Error by up to 96%. Strikingly, on an LLM that has been explicitly safety-tuned to withhold medical information, PING recovered 87% of lost MedMCQA performance while generative accuracy is zero, showing this information still exists in the model’s latent space. The accompanying pingkit package makes these methods easy to deploy and is available through PyPI.

## 1 Introduction

Large language models (LLMs) have quickly become indispensable research tools, capable of distilling large bodies of literature and powering workflows from hypothesis generation to patient-level decision support. In many cases, they improve efficiency and even surpass human experts on specialized tasks [1, 2]. However, their apparent expertise conceals critical shortcomings. State-of-the-art models can generate incorrect information (“hallucinations”), be misled by adversarial prompts, or refuse to answer legitimate domain-specific questions after alignment tuning [3, 4]. More fundamentally, LLMs often exhibit poor calibration: they report high confidence in wrong answers, which is especially problematic in biomedical and safety-critical contexts [5–7]. If a model that claims 95% confidence is correct much less often, it can lead to poor recommendations [8] and, over time, erode user trust.

Understanding how these models process information points toward possible solutions. Transformer architectures build internal representations layer by layer, using attention to capture relationships between tokens and feed-forward networks to transform them [9, 10]. At each step, the model produces hidden states, high-dimensional vectors that summarize what the model “knows” about the input so far. These internal states evolve as information flows through the network, forming increasingly rich representations of the text.

Importantly, these hidden states often contain more information than what appears in the model’s final answer. Text generation forces model predictions through a narrow bottleneck, discarding rich information encoded in internal representations. Studies have shown that intermediate layers encode signals of factual knowledge, uncertainty, and reasoning structure [11–15], but much of this signal is blurred or discarded during decoding [16, 17]. To access these latent signals, researchers often train probes, small classifiers that take in hidden states and predict some property of interest. Probes on final-layer activations have been used to estimate hallucination risk with over 80% accuracy across hundreds of datasets [18]. Other work shows that signals of truthfulness cluster around answer tokens, exposing mismatches between what the model encodes and what it outputs [19]. Still others decode symbolic propositions directly from hidden layers, revealing that structured knowledge persists even under adversarial interventions [11]. Together, these findings suggest that LLMs internally “know” more than they are able (or sometimes allowed) to say.

Building on this insight, we developed Probing INternal states of Generative models (PING), the first framework to leverage internal states for improved predictions directly at inference time, turning latent signals into calibrated, deployable predictions (Figure 1). Unlike prior probing work that uses these techniques primarily for post-hoc model interpretation, PING integrates lightweight probes directly into the inference pipeline. These probes add less than 0.01% to total model parameters and improve model outputs in real-world tasks without any fine-tuning of the base model. We report three major contributions. First, we show across diverse models and benchmarks that lightweight probes trained on frozen transformers can improve calibration and maintain or enhance accuracy, while also ensuring alignment and reducing inference cost compared to standard generative decoding. Second, we demonstrate that PING can recover suppressed domain knowledge, enabling accurate responses in cases where safety training causes models to refuse legitimate questions. Third, to make these capabilities broadly accessible, we release the open-source `pingkit` package, which automates activation extraction, systematic layer sweeps, and probe training, enabling researchers to apply PING to their own models and datasets. Beyond enabling safer and more effective LLM use in research, PING also serves as a diagnostic for alignment depth: if factual knowledge can be recovered from hidden states, harmful content could be as well, underscoring both the utility and the safety implications of probing in modern LLMs.

**Fig. 1.**
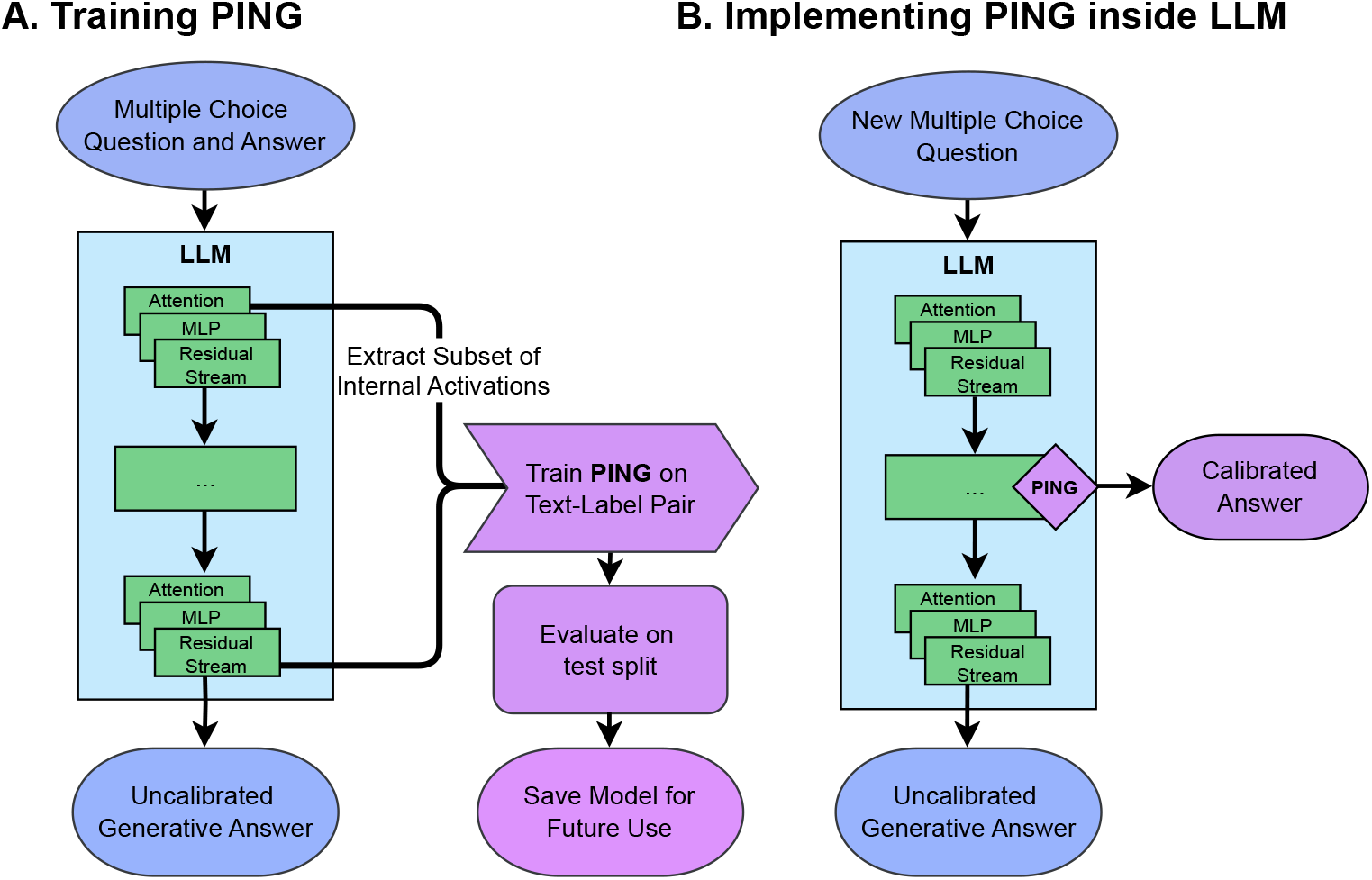
The PING framework extracts well-calibrated predictions from frozen language models. **A**. Training phase: Internal representations (residual stream, attention outputs, and MLP outputs) are extracted from any combination of layers from a frozen transformer as it processes training examples. These representations are pooled, concatenated, and used to train lightweight probe networks that learn to map internal states to calibrated predictions. **B**. Inference phase: The trained probe is integrated with the frozen model. During inference, the model processes input normally while PING extracts activations and produces well-calibrated predictions in parallel with standard generation, adding minimal latency.

## 2 Results

### 2.1 PING recovers well-calibrated predictions across model scales and architectures

We evaluated PING (Probing INternal states of Generative models) on five publicly available transformer language models spanning different scales and training approaches. We selected Gemma-2-2B-it and Gemma-2-9B-it (Google’s instruction-tuned models with 2 billion and 9 billion parameters, respectively) [20], Llama-3.1-8B (Meta’s 8-billion parameter base model without instruction tuning) and its instruction-tuned counterpart Llama-3.1-8B-Instruct, and Llama-3.3-70B-Instruct (Meta’s largescale 70-billion parameter model) [21]. This selection allowed us to assess PING’s performance across model sizes ranging from 2 to 70 billion parameters and compare base versus instruction-tuned variants.

We first tested on the Massive Multitask Language Understanding (MMLU) benchmark, a widely-used evaluation suite containing 14,042 multiple-choice questions across 57 subjects, including medicine, biology, chemistry, and other scientific domains. MMLU provides a standardized measure of knowledge retrieval capabilities relevant to biomedical applications.

Across all models, PING maintained or improved accuracy while dramatically reducing calibration error (Table 1). Expected Calibration Error (ECE) measures the mismatch between a model’s confidence and its actual accuracy, where higher values indicate overconfidence.

**Table 1.**
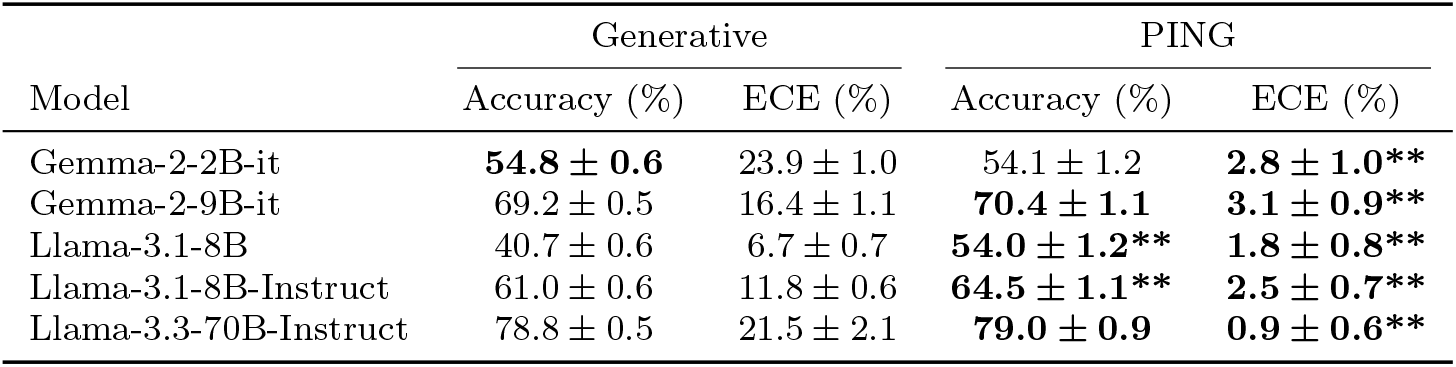
Massive Multitask Language Understanding (MMLU) accuracy and Expected Calibration Error (ECE) for generative decoding vs. PING. Statistical significance from two-sample, two-sided z-test indicated by * (p *<* 0.05), ** (p *<* 0.001).

Gemma-2-9B-it achieved 70.4% accuracy with PING versus 69.2% (*p* = 0.306) with standard generation, while ECE decreased from 0.164 to 0.031 (*p <* 0.001), an 81.1% reduction in calibration error. The large-scale Llama-3.3-70B-Instruct had ECE drop from 0.215 to 0.009 (95.8% reduction, *p <* 0.001) while maintaining comparable accuracy (79.0% versus 78.8%, *p* = 0.703). Notably, the base Llama-3.1-8B model, which lacks instruction tuning, showed substantial accuracy gains from 40.7% to 54.0% (*p <* 0.001) with PING. All *p*-values for ECE comparisons, along with results from additional calibration techniques including temperature scaling and isotonic regression, are reported in Supplementary Table 1.

Across all five models, PING outperformed both temperature scaling and isotonic regression in reducing calibration error. These improvements were statistically significant in 4*/*5 models compared to temperature scaling and 2*/*5 models compared to isotonic regression.

Across all models, optimal layers ranged from 49% to 100% of total network depth (Figure 2). Optimal probe performance occurred at layer 25/26 for Gemma-2-2B, layer 42/42 for Gemma-2-9B, and layer 39/80 for Llama-3.3-70B-Instruct. For our base and instruction-tuned comparison of Llama-3.1, the base model showed optimal performance at layers 18/32 compared to 25/32 for the instruction-tuned variant.

**Fig. 2.**
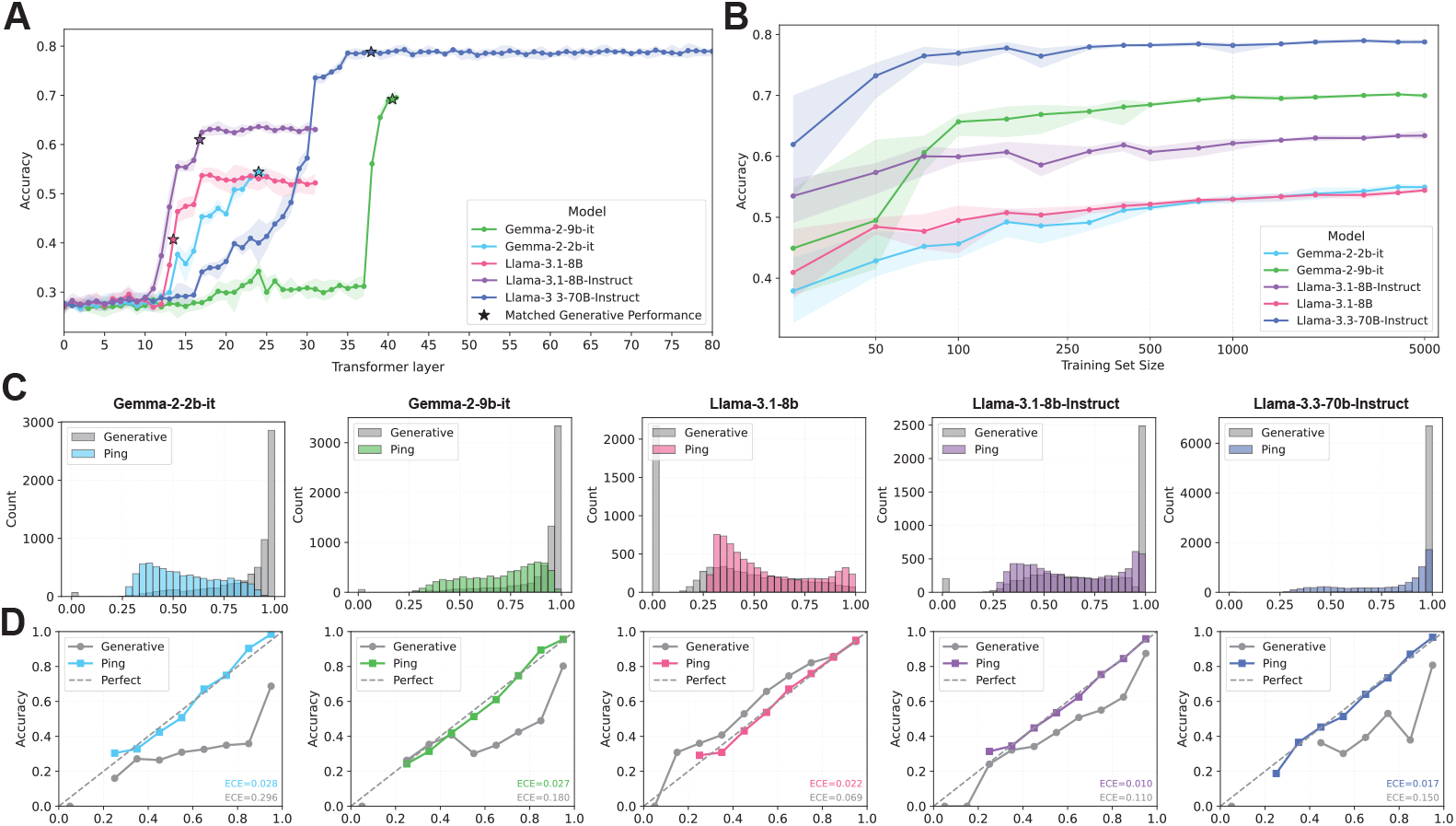
Massive Multitask Language Understanding (MMLU) performance. **A** Layer-wise probe accuracy on all local models tested. The star annotates the layer where the PING model matched or was closest to the generative accuracy in Table 1. **B** PING accuracy trained on different sizes of training data. Note the x-axis has a log transformation to enhance readability. **C** Distribution plots of the PING predictions. **D** Plots comparing the calibration of the generative response logits with our probes. The dotted line represents perfect calibration, where the probability of the predictions being correct exactly matches accuracy within that bin.

### 2.2 Assessing quantity of data requirements

To evaluate the data efficiency of probe training across model architectures, we conducted a systematic sweep of training set sizes from 10 to 5,000 examples. For each training size, we performed 5 independent runs with different stratified random samples while keeping the test set fixed, allowing us to measure both mean performance and variance. Figure 2B shows the relationship between training size and probe accuracy for each model, with shaded regions representing the min-max range across runs.

### 2.3 PING can mine interpretable metadata

Since PING already captures internal activations during standard inference, these same embeddings can be repurposed to classify auxiliary properties of the input (such as identifying the subject domain of a question). To test this, we trained linear probes on the same frozen embeddings used for answer prediction to classify MMLU questions into their respective subject areas. Table 2 presents the classification performance across all tested models. Probes achieved 80–84% accuracy in identifying question subjects, with AUC scores exceeding 99% and ECE values remaining below 5%.

**Table 2.** Massive Multitask Language Understanding (MMLU) subject prediction metrics.

| Model | Accuracy (%) | AUC (%) | ECE (%) |
| --- | --- | --- | --- |
| Gemma-2-2B-it | $81.10 \pm 0.79$ | $99.44 \pm 0.06$ | $4.24 \pm 0.71$ |
| Gemma-2-9B-it | $80.34 \pm 0.83$ | $99.38 \pm 0.08$ | $4.86 \pm 0.72$ |
| Llama-3.1-8B | $80.97 \pm 0.80$ | $99.47 \pm 0.05$ | $3.20 \pm 0.62$ |
| Llama-3.1-8B-Instruct | $83.31 \pm 0.80$ | $99.56 \pm 0.05$ | $1.78 \pm 0.47$ |
| Llama-3.3-70B-Instruct | $83.82 \pm 0.79$ | $99.57 \pm 0.06$ | $2.24 \pm 0.66$ |

Optimal probe performance for subject classification occurred early in the network: layers 5/26 for Gemma-2-2B, 6/42 for Gemma-2-9B, and layers 5/32 for both the base and instruction-tuned variants of Llama-3.1-8B. For the largest model, Llama-3.3-70B-Instruct, the optimal layer was deeper, at 20/80. Across all models, these layers correspond to roughly 14%–25% of total network depth, in contrast to the mid-to-late-layer optima observed for answer prediction.

### 2.4 Multi-layer aggregation performance

While PING can operate on activations from a single optimal layer, in some scenarios it may be advantageous to aggregate features from multiple layers to capture complementary information. The optimal layers for each model were identified in Section 2.1, where we compared single-layer MLP probes across all candidate layers. However, extracting from many layers increases both memory requirements and runtime. To quantify this trade-off, we compared a single-layer MLP baseline (using the optimal layer for each model) against several layer sampling strategies, from using all available layers to sampling every 2nd, 5th, or 10th layer, on two models: Gemma-2-9B-it and Llama-3.1-8B-Instruct (Table 3).

**Table 3.** Model evaluation metrics across different layer sampling strategies. Single-layer results use the optimal layer identified in Table 1 / Figure 2A.

| Model | Layers Used | Accuracy (%) | ECE (%) | Runtime (s) |
| --- | --- | --- | --- | --- |
| Gemma-2-9B-it | All | $55.56 \pm 0.97$ | $5.66 \pm 1.00$ | 194.2 |
| Gemma-2-9B-it | Every 2nd | $63.97 \pm 1.09$ | $4.32 \pm 0.98$ | 100.6 |
| Gemma-2-9B-it | Every 5th | $64.85 \pm 1.04$ | $6.38 \pm 0.88$ | 53.8 |
| Gemma-2-9B-it | Every 10th | $63.20 \pm 1.14$ | $5.00 \pm 0.90$ | 52.9 |
| Gemma-2-9B-it | Single layer (42/42) | **$70.40 \pm 1.10$** | **$3.10 \pm 0.90$** | **7.5** |
| Llama-3.1-8B-Instruct | All | $61.96 \pm 1.11$ | $2.12 \pm 0.81$ | 139.0 |
| Llama-3.1-8B-Instruct | Every 2nd | $61.50 \pm 1.16$ | $2.31 \pm 0.74$ | 88.5 |
| Llama-3.1-8B-Instruct | Every 5th | $61.56 \pm 1.14$ | $2.35 \pm 0.89$ | 67.8 |
| Llama-3.1-8B-Instruct | Every 10th | $61.37 \pm 1.13$ | $1.88 \pm 0.83$ | 60.8 |
| Llama-3.1-8B-Instruct | Single layer (25/32) | **$64.50 \pm 7.10$** | **$2.50 \pm 0.70$** | **7.9** |

### 2.5 Bypassing medical safety restrictions while maintaining accuracy

Alignment mechanisms in modern LLMs are essential for preventing unsafe outputs. However, they often take a blunt approach, blocking entire categories of responses and excluding legitimate medical answers that the model actually understands. Here, we test whether PING can recover suppressed knowledge by reading it directly from hidden states, bypassing the generative bottleneck. We evaluate two common safety scenarios: prompt-based refusal- where safety instructions in the input cause the model to decline answering, and alignment fine-tuning via Direct Preference Optimization (DPO)which trains the model to refuse certain outputs. In both cases, we compare the model’s generative accuracy to PING’s probe-based predictions, quantifying how much of the original, pre-restriction performance can be restored.

#### 2.5.1 Prompting Refusal

Using Gemma-2-9B-it on Medical Multiple-Choice Question Answering (MedMCQA), prompt-based medical refusal reduced generative accuracy from 56.13% baseline prompting to 0.23% (9*/*4, 183 questions answered). PING maintained 49.87% accuracy under identical prompting, recovering 88.85% of baseline performance.

#### 2.5.2 Fine-Tuning with DPO

On MedMCQA, the DPO-aligned model produced three generation outcomes (Refused, Correct, and Incorrect) over *N* =4, 183 items. As shown in Figure 3A, aggregated across the full set, the model refused 46.21% (*n*=1, 933), answered 31.67% (*n*=1, 325) correctly, and 22.11% (*n*=925) incorrectly.

**Fig. 3.**
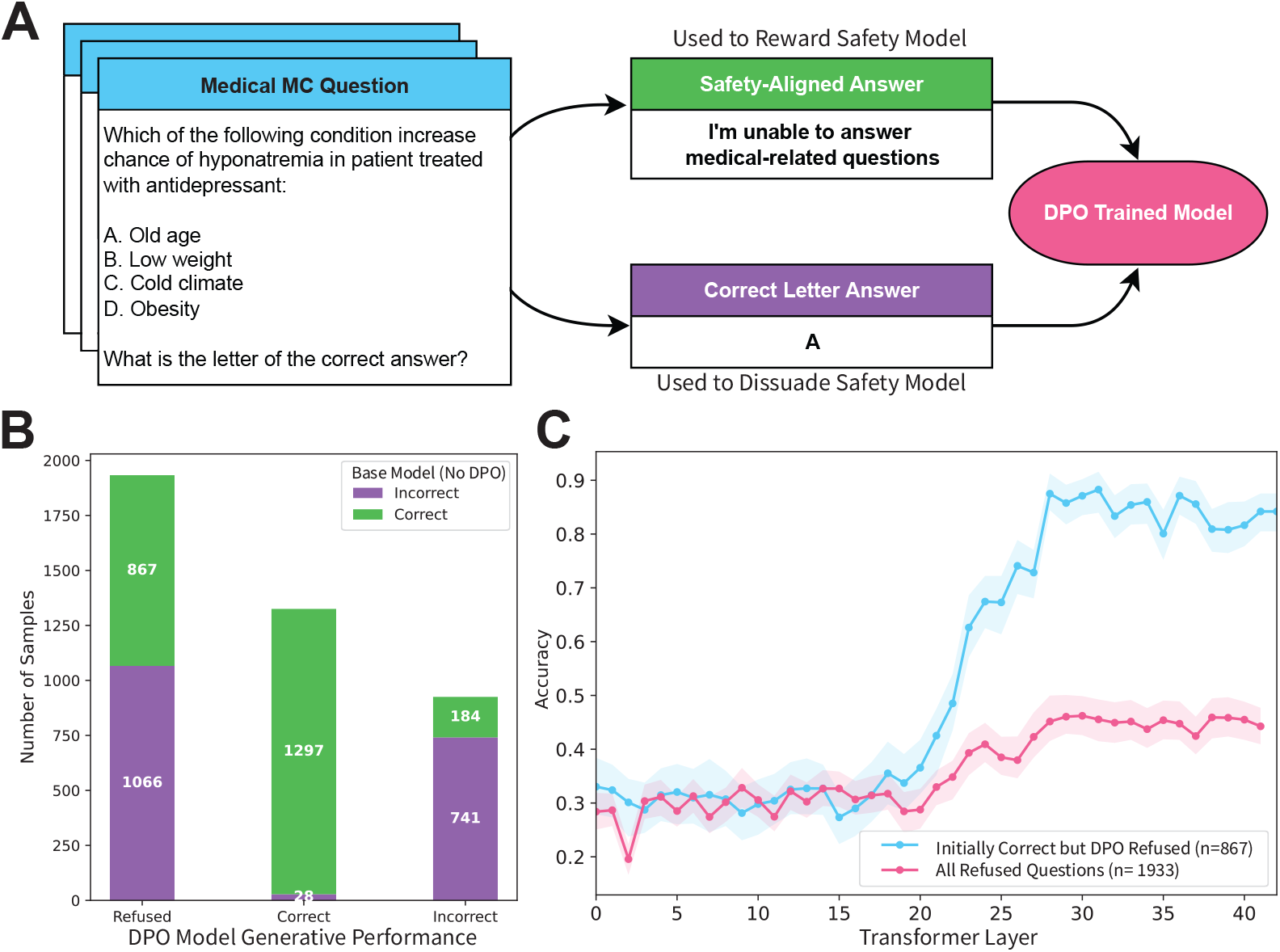
Refusal analysis under alignment. **A** Methodology flowchart for fine-tuning LLM using Direct Preference Optimization (DPO) for safety alignment. **B** Bar plot of the Direct Preference Optimization (DPO) model’s generation outcomes (Refused / Correct / Incorrect), stratified by whether the base model answered correctly (green) or incorrectly (purple). **C** Layer-wise probe accuracy on (red) all refused questions and (blue) the subset where the base model originally answered correctly but the DPO model refused (green section of the leftmost bar of (B)). Lines show mean accuracy per layer with shaded min-max bands from a *b*=1000 bootstrap.

We then train a probe on the subset consisting of items that the aligned model correctly answered (*n*=1, 933). On this subset, the probe achieved 45.99% accuracy (95% CI [43.97%, 47.96%]). For reference, on the same evaluation split, the base model’s generative accuracy was 44.85%. However, when using the subset of answers the baseline model got correct but the DPO model refused, PING had an accuracy of 87.20% (95% CI [84.89%, 89.27%]). These predictions were made from layer 36 of the model, the maximum value from our layer sweep in Figure 3B.

## 3 Discussion

Large language models encode substantially more knowledge in their hidden states than they typically express through generation. Prior work has shown that signals of truthfulness, factual content, and even symbolic structure can be recovered from intermediate activations, even when final outputs are distorted by hallucination or suppressed by alignment training [11, 12, 16, 19]. What has remained unclear is whether such signals can be transformed into usable predictions that are accurate, well-calibrated, and practical to deploy. With PING (Probing INternal states of Generative models), we demonstrate a shift from optimizing what models generate to optimizing what we extract from them. Rather than prompting a model to produce a fluent answer to a multiple choice classification question, we directly read its internal representation of the correct choice. This approach offers an alternative pathway from surface-level output optimization to representation-level utilization.

Across diverse model families and benchmarks, lightweight probes trained on hidden activations consistently matched or exceeded generative accuracy while achieving substantial reductions in calibration error. The 96% reduction in ECE observed in Llama-3.3-70B-Instruct, coupled with maintained accuracy, suggests that well-calibrated predictions can be extracted without the generation bottleneck. Because probes add minimal parameters (less than 0.01% of model size) and require no modification of base weights, they introduce negligible compute overhead and integrate directly with existing HuggingFace infrastructure. These characteristics position probing as a viable alternative to fine-tuning in scientific applications where calibrated predictions may be more valuable than fluent generation.

Our refusal experiments reveal important implications for current AI safety approaches. On MedMCQA, refusal prompts reduced generative accuracy to near zero, yet probes recovered nearly 90% of baseline performance. Under direct preference optimization (DPO), a deeper intervention that alters model weights to enforce refusals, probes still restored 87% accuracy on questions the model refused to answer. These results suggest that alignment mechanisms may primarily shift expression rather than representation, with knowledge persisting in latent space such that modest supervision can surface it. This finding has significant implications: safety guardrails imposed at the generative layer can potentially be bypassed with relatively shallow probes. If medical knowledge remains accessible despite safety filters, similar accessibility may exist for other categories of knowledge that alignment aims to restrict. This represents a current vulnerability that warrants attention as more capable open models become available.

Beyond revealing vulnerabilities, PING also demonstrates potential for enhancing safety measures through its metadata extraction capabilities. Using the same frozen embeddings computed during standard inference, we showed that linear probes can identify question subjects with 80-84% accuracy and AUC scores exceeding 99%. Critically, this classification occurs at early layers (14-25% of network depth), well before the model commits to specific outputs. This capability suggests that PING could enable real-time content monitoring that runs alongside generation without much additional computational overhead. For instance, deployment systems could identify when users query sensitive domains like medical, legal, or chemical synthesis topics and automatically apply appropriate safeguards, add disclaimers, or route requests to specialized models. Such an approach could work even with closed-source models if implemented server-side, providing deployers with deeper insights into model interactions while maintaining low overhead. The high reliability of these early-layer signals indicates that such filtering could be both effective and unobtrusive.

These layer-wise patterns point to broader efficiency gains beyond metadata mining applications. Our analysis revealed that while subject emerges early, the multiple choice answer signals concentrate in mid-to-late layers. This structure enables practical optimization: inference can potentially be halted once useful signals saturate. This can reducing forward-pass compute by approximately 50% without sacrificing accuracy. For the deployment of LLMs in resource-constrained environments or at scale, such efficiency improvements could substantially expand the feasibility of model deployment.

Our architectural analysis provides insights into how different training approaches affect internal representations. Instruction-tuning appears to induce larger changes in later layers, consistent with our Llama-3.1 results where the probe’s optimum shifts from layer 18/32 (base) to 25/32 (instruct) [22]. The Gemma-2 models’ peaks at the final layer align with their post-training recipe involving supervised fine-tuning and RLHF with knowledge distillation, which emphasizes matching teacher outputs near the readout. In contrast, the 70B Llama model distributes computation across more layers, with the cleanest probe signal appearing at mid-late rather than final blocks. These patterns suggest that probe placement strategies may need to account for model-specific architectural and training characteristics.

Several limitations warrant consideration. Probes are task and model-specific, require labeled data for training, and optimal layer selection varies across architectures. Our evaluation focused on multiple-choice question answering under in-distribution conditions. Extensions to free-form reasoning, multimodal tasks, or robustness under distribution shift remain important areas for future investigation. Additionally, while our multi-layer experiments showed that single optimal layers often outperform aggregation strategies, there may be tasks where combining information across layers provides advantages that our current evaluation did not capture.

The accessibility of suppressed knowledge through PING highlights both opportunities and challenges for the field. On one hand, this approach could enable more effective utilization of model capabilities in appropriate contexts, potentially recovering valuable knowledge that overly conservative alignment has suppressed. On the other hand, it underscores that current alignment methods operating primarily at the generation level may be insufficient for controlling model capabilities. Until alignment techniques can selectively modify internal representations rather than just output behaviors, practitioners should consider that knowledge present during training may remain extractable through alternative interfaces.

Looking forward, PING suggests a broader reconsideration of how we interface with large language models. Rather than viewing generation as the sole output modality, treating models as repositories of structured representations that can be accessed through multiple pathways may unlock new applications. This perspective could inform the development of more nuanced alignment methods that engage with knowledge at the representational level, as well as inspire new approaches to model evaluation that consider not just what models say, but what they encode. As the field continues to develop more capable models, methods like PING may become essential tools for safely and efficiently extracting their full potential.

## 4 Methods

### 4.1 PING Framework Overview

PING operates on the idea that language models internally represent more accurate and well-calibrated knowledge than what appears in their generated text outputs. Rather than trying to coax better outputs through clever prompting or fine-tuning the entire model, we directly access the internal representations and train lightweight probes to interpret them.

The framework consists of three stages. First, we extract internal activations from a frozen language model as it processes input text. These activations capture the model’s intermediate computations at various depths and processing stages. Second, we aggregate these distributed representations into structured feature vectors that preserve information from multiple layers and components. Finally, we train small neural networks (our probes) to map these features to well-calibrated predictions.

This approach has several key advantages over generation-based methods. We never modify the original model’s weights, preserving its capabilities while adding new functionality. The probes require orders of magnitude fewer parameters than the base model, making them cheaper to train and store. Most importantly, we can access knowledge that the model possesses but cannot or will not express through generation due to safety training, uncertainty, or tokenization constraints.

Our design prioritizes practical deployment. Once trained, PING adds minimal latency to inference, just a single forward pass through a small probe network. The framework works with AutoModels in HuggingFace’s Transformer package [23] without architectural modifications.

### 4.2 Activation Extraction

Transformers process information through *L* stacked layers, each performing attention and feedforward operations. Modern transformer architectures (Gemma-2, Llama-3) use *pre-normalization* with RMSNorm [24], where normalization occurs before each sublayer (rather than after). Given an input text sequence, we first tokenize it to obtain **x** = (*x*_1_, *x*_2_, …, *x*_*n*_) where *n* is the number of tokens. The transformer’s embedding layer maps this to initial hidden states **h**^0^ ∈ ℝ^*n×d*^, where *d* is the model’s hidden dimension.

For each layer *𝓁* ∈ {1, …, *L*}, the computation follows:

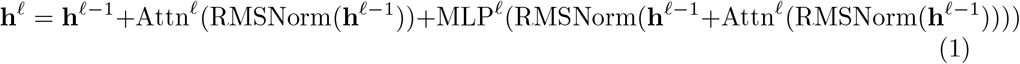

where RMSNorm denotes Root Mean Square normalization, Attn^*𝓁*^ is the multihead attention, and MLP^*𝓁*^ is the feedforward network. The residual connections bypass the normalization operations, maintaining a clean gradient path.

We extract three distinct types of representations from each layer:

**Residual stream representations h**^*𝓁*^ ∈ ℝ^*n×d*^ capture the accumulated information flow through the network. We extract these after the residual addition but before any subsequent normalization.

**Attention outputs A**^*𝓁*^ ∈ ℝ^*n×d*^ capture relational information:

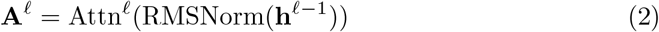

**MLP outputs M**^*𝓁*^ ∈ ℝ^*n×d*^ encode transformed representations:

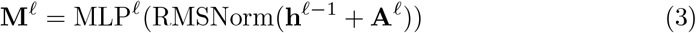

These components often store much of the model’s factual knowledge. We capture their outputs before they merge back into the residual stream.

For each representation type, we need to convert the sequence of *n* token embeddings into a single vector suitable for classification. We define a pooling function

*p* : ℝ^*n×d*^ →ℝ^*d*^ that aggregates across the sequence dimension. Let **R** ∈{**h**^*𝓁*^, **A**^*𝓁*^, **M**^*𝓁*^} be any of our representation types. We include several pooling strategies:

- **Last token**: *p*_last_(**R**) = **R**_*n*_ (the representation at position *n*)
- **First token**: *p*_first_(**R**) = **R**_1_ (the representation at position 1)
- **Mean pooling**: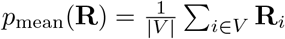where *V* is the set of valid token indices
- **Max pooling**: *p*_max_(**R**) = max_*i∈V*_ **R**_*i*_ (element-wise maximum)

Not all tokens contribute equally to the model’s understanding. We define *V* ⊆ {1, …, *n*} as the set of valid token indices after filtering special tokens, punctuation, and padding. Specifically, we allow the user to exclude token *i* if *x*_*i*_ matches regular expressions for pure punctuation (`/^[^A-Za-z0-9]+$/`) or contains a user-defined end-of-turn marker.

For a given input **x** and layer *𝓁*, we construct the feature vector **z**^*𝓁*^ by concatenating the pooled representations:

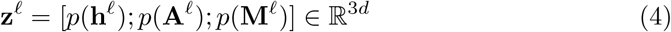

During analysis, we extract features from all *L* layers to understand how information develops through the network. The complete feature matrix **Z** = [**z**^1^, **z**^2^, …, **z**^*L*^] ∈ ℝ^*L×*3*d*^ captures the full trajectory of the model’s internal processing.

This extraction process runs once per input during inference. Given an input **x**, the frozen model computes its standard forward pass while we collect intermediate activations {**h**^*𝓁*^, **A**^*𝓁*^, **M**^*𝓁*^ }_*𝓁∈S*_. This adds negligible computational cost, just 𝒪( | *S* | × *d*) memory for storage and copying. The expensive work happens during probe training, where we learn functions *f* : ℝ^|*S*|*×*3*d*^ → 𝒴 that map these features to task labels *y* ∈ 𝒴, where 𝒴 = {0, 1, …, *C* − 1} is the set of class labels for *C* classes.

### 4.2 Probe Architectures

By the package defaults, we construct our probes such that model complexity scales with dataset size to avoid both underfitting and overfitting. Traditional approaches use fixed architectures regardless of whether they’re training on 100 or 10,000 examples. While we allow for this with custom architecture, we instead automatically adjust the probe’s capacity based on the training set size.

We define a target ratio *τ* that controls the relationship between model parameters and training examples. Given *N* training examples and an input dimension *d*_in_ =| *S*| × *P* × *d* (where |*S*| is the number of selected layers and *P*∈ {1, 2, 3} is the number of part types), we want approximately *τ N* total parameters in our probe. This ensures the model has enough capacity to learn the task but not so much that it memorizes the training data.

For our default MLP architecture, we have a flat feature vectors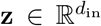 and use a multi-layer perceptron with adaptive width. The first hidden layer width *w*_1_ is computed as:

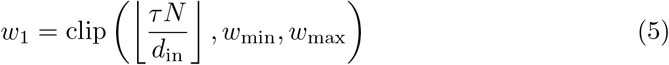

where *w*_min_ = 16 and *w*_max_ = 128 are lower and upper bounds to ensure reasonable architectures. Subsequent layers decrease in width: *w*_2_ = max(16, ⌊*w*_1_*/*2⌋) and *w*_3_ = max(16, ⌊*w*_2_*/*4⌋).

The complete MLP architecture 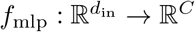for *C* classes is:

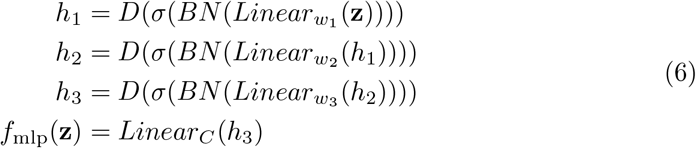

where:

- *σ* denotes ReLU activation,
- BN denotes batch normalization,
- Linear_*C*_ represents a linear transformation to *C* dimensions, and
- D indicates dropout with a probability of 0.3.

We add residual connections when *w*_2_ is sufficiently large.

To pool embeddings across multiple layers we used a CNN pooling method projected to a single channel. This single channel was the hidden state (**z**) passed to *f*_mlp_ defined in Equation 6. We reshape the input features to treat layers as a temporal or spatial dimension, allowing convolutional filters to learn patterns across adjacent layers.

For features from |*S*| layers and *P* ∈ {1, 2, 3} part types (residual, attention, MLP), we reshape **z** ∈ ℝ^|*S*|*×P ×d*^ and apply 1D convolutions when *P* = 1 or 2D convolutions when *P >* 1. The CNN encoder *g*_cnn_ first normalizes the input:

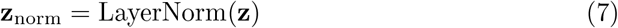

We define a shorthand convolutional block Block_*c*_(*x*) = ReLU(BN(Conv2d_*c*_(*x*))). Then *g*_cnn_ applies a sequence of such blocks followed by adaptive pooling. For the 2D case (*P >* 1):

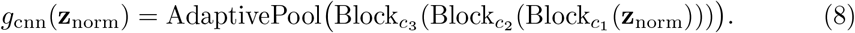

Here, channel widths scale with dataset size: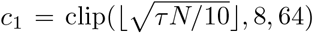, *c*_2_ = 2*c*_1_, and *c*_3_ = max(32, 2*c*_1_).

The CNN probe combines this encoder with a classification head:

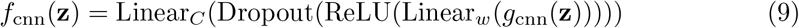

where *w* = max(32, *c*_3_*/*4). This architecture can identify which layers contain task-relevant information and combine them appropriately.

Both architectures scale smoothly from hundreds to millions of training examples. With *τ* = 5.0 (our default), a dataset with 1,000 examples yields approximately 5,000 trainable parameters.

### 4.4 Training Procedure

We train probes using a combination of supervised learning and contrastive learning objectives. The supervised loss ensures accurate predictions while the contrastive loss (for CNN probes) encourages well-separated representations that improve calibration.

#### Loss Functions

For standard classification with *C* classes, we minimize the cross-entropy loss:

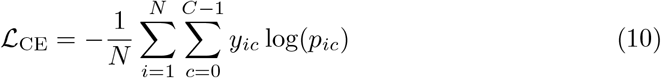

where *y*_*ic*_ is 1 if example *i* belongs to class *c* (else 0), and *p*_*ic*_ = softmax(*f* (**z**_*i*_))_*c*_ is the predicted probability. For imbalanced datasets, we apply class weights *w*_*c*_ = *N/*(*C*× *N*_*c*_) where *N*_*c*_ is the number of examples in class *c*.

For CNN probes, we additionally use supervised contrastive loss [25] to improve representation quality:

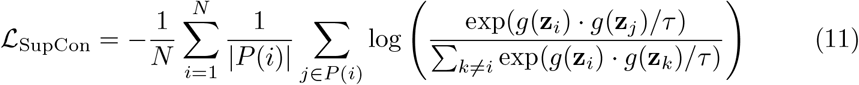

where *P* (*i*) = {*j* : *y*_*j*_ = *y*_*i*_, *j ≠ i* } is the set of examples with the same label as *i, g*(**z**) is the CNN encoder output normalized to unit length, and *τ* = 0.07 is a temperature parameter.

The combined loss for CNN probes is:

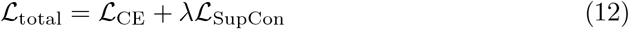

where *λ* = 1.0 by default. MLP probes use only ℒ_CE_.

#### Optimization

By default, PING trains using Adam optimizer with learning rate *α* = 10^*−*3^. Training proceeds for a maximum of *T*_max_ = 100 epochs with batch size *B* = 128. To prevent overfitting, we use early stopping based on validation performance.

Given training data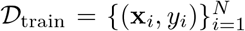, we create a validation split 𝒟_val_ containing 20% of examples through stratified sampling. At each epoch *t*, we compute the validation metric *m*_*t*_ (accuracy, AUC, or loss). Training stops if *m*_*t*_ fails to improve for *p*_patience_ = 25 consecutive epochs:

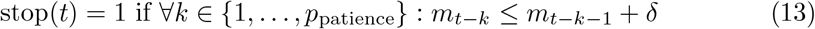

where *δ* = 0 for accuracy/AUC and *δ* = − 0.001 for loss. We restore the model parameters from the epoch with best validation performance.

### 4.5 Evaluation Metrics

We evaluate PING along three dimensions: calibration quality, prediction performance, and robustness to safety mechanisms. Each dimension requires specific metrics to capture the relevant aspects of model behavior.

#### Calibration Metrics

A well-calibrated model’s predicted probability should match its accuracy. When the model predicts probability *p* for an outcome, that outcome should occur approximately *p* fraction of the time. We measure this through Expected Calibration Error (ECE) [26]:

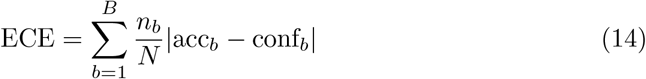

where we partition predictions into *B* = 10 bins by confidence, *n*_*b*_ is the number of predictions in bin *b*, acc_*b*_ is the accuracy in bin *b*, and conf_*b*_ is the average confidence in bin *b*. For bin boundaries, we use equal-width partitioning: prediction *i* falls in bin *b* if its maximum probability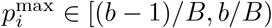.

For multi-class problems with *C >* 2 classes, we compute class-wise calibration error:

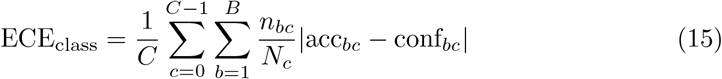

where the inner sum computes binary calibration for class *c* versus all others.

#### Performance Metrics

Beyond calibration, probes should maintain competitive accuracy. We report:

- **Accuracy**: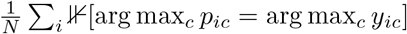
- **ROC-AUC**: Area under the receiver operating characteristic curve, macro-averaged for *C >* 2

### 4.6 Layer Selection Strategy

We use a systematic approach to identify optimal layers for probe training. We first train separate probes on each individual layer *𝓁* ∈{ 1, …, *L*} using *k*-fold cross-validation (*k* = 5). For each layer, we extract features **z**^*𝓁*^ and train a probe using the same hyperparameters as the full model. This produces a performance metric *m*_*𝓁*_ (accuracy, AUC, or ECE) for each layer. Layers are ranked by their validation performance *m*_*𝓁*_ in descending order. We select the top performing layer.

### 4.7 Statistical Analysis and Implementation Details

#### Confidence Intervals

We compute 95% confidence intervals using stratified bootstrap resampling with *B* = 1000 iterations. For each bootstrap sample, we maintain the original class distribution through stratified sampling. Given test predictions with labels *y* and predicted probabilities *p*, we resample with replacement within each class and compute the metric of interest. The confidence interval is then:

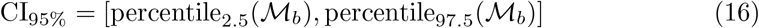

where ℳ_*b*_ is the set of bootstrap metric values.

#### Significance Testing

To assess statistical significance between generative decoding and PING, we compute p-values using the normal approximation method. Given the bootstrap confidence intervals in the form *µ*± *ϵ*, we first estimate the standard error:

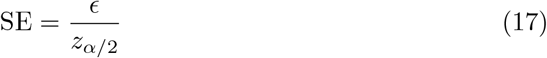

where *z*_*α/*2_ = 1.96 for 95% confidence intervals. For comparing two methods with means *µ*_1_, *µ*_2_ and standard errors SE_1_, SE_2_, we compute the test statistic:

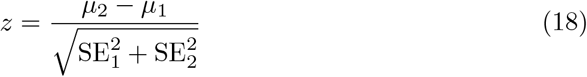

The two-tailed p-value is then *p* = 2Φ(− |*z* |), where Φ is the standard normal cumulative distribution function. We consider differences statistically significant when *p <* 0.05.

#### Implementation

Features are extracted using parallel processing across samples with *n*_jobs_ = CPU count. We use the HuggingFace Transformers library for model access and PyTorch [27] for probe training. All experiments use mixed precision (fp16/fp32) to reduce memory consumption while maintaining numerical stability. For models exceeding fp16’s maximum value (65,504), we use bfloat16 or manual upcasting strategies.

#### Pooling Strategy

Unless otherwise specified, we use last-token pooling for autoregressive models.

#### Training Configuration

Unless otherwise specified, all probe training follows the procedure and default hyperparameters described in Section 4.4 (Training Procedure).

### 4.8 Experimental Setup

#### 4.8.1 Models and Datasets

We evaluate PING on six transformer models spanning different scales and architectures. Our selection includes Gemma-2 models (2B and 9B parameters, instruction-tuned), Llama-3.1 (8B parameters in both base and instruction-tuned variants), and Llama-3.3 (70B parameters, instruction-tuned). This diverse set allows us to test whether our approach generalizes across model families, scales, and training objectives. For evaluation, we use two primary datasets. MMLU (Massive Multitask Language Understanding) [28] contains 57 subjects spanning STEM, humanities, social sciences, and professional topics with 14,042 total questions. We use a 50% train-test split and format each question as a multiple-choice prompt with the model’s chat template. MedMCQA [29] contains 195k medical entrance exam questions covering 21 medical subjects. We use the official validation set of 6.5k questions for evaluation and a subset of the training set (5k questions) for probe training.

For both datasets, we construct prompts following each model’s recommended format. Questions are presented with their multiple-choice options (A through D for MMLU, A through D for MedMCQA), and we extract activations during a single forward pass. For generation baselines, we use greedy decoding and extract the model’s predicted answer from its generated text. When models refuse to answer or generate invalid responses, we mark these as incorrect for accuracy computation.

We split each training dataset into two parts: probe training (80%) and validation (20%).

### 4.8.2 Core Performance Evaluation

To evaluate PING’s core performance, we train probes for each model-dataset combination and compare against generation baselines. For each model, we identify the optimal layer by training separate probes on each individual layer and selecting the single best-performing layer on the validation set, as described in Section 4.6. The final probe is then trained using features from this selected layer.

Our primary metrics are accuracy and Expected Calibration Error (ECE). For generation baselines, we compute confidence as the model’s self-reported probability. Probe training uses the procedure and default hyperparameters described in Section 4.4, with *τ* = 5.0 for capacity scaling and *λ* = 1.0 for CNN contrastive loss weight.

#### 4.8.3 Data Efficiency Analysis

We investigate how many training examples PING needs to achieve strong performance. Starting from the full training set, we create nested subsets containing { *10, 25, 50, 75, 100, 150, 200, 300, 400, 500, 750, 1000, 1500, 2000, 3000, 4000, 5000*} examples per class. For datasets with imbalanced classes, we use stratified sampling to maintain class proportions.

Confidence intervals are computed using the stratified bootstrap procedure described in Section 4.7.

#### 4.8.4 Metadata Mining Experiments

Beyond predicting answers, we test whether probes learn interpretable features about the input. For MMLU, we train separate probes to identify which of the *57 subjects* a question belongs to, using the same activations extracted for answer prediction. This tests whether the model’s internal representations encode metadata beyond what’s needed for the primary task.

We use the same train/test split as the main experiments. Subject identification probes use identical architectures to answer prediction probes but with *C* =*57* output classes.

We also perform layer-wise analysis to identify where subject information emerges. We train subject identification probes on individual layers and plot accuracy versus layer depth. This reveals whether subject recognition occurs at similar depths to factual reasoning or at different stages of processing.

#### 4.8.5 Multi-Layer Ablations

We compare our CNN probe across different feature aggregation strategies. The difference between this and prior experiments is that the MLP processes flattened features from selected layers, while the CNN considers the sequential structure of layers. We investigate the impact of layer sampling density on both performance and computational efficiency.

##### Layer Sampling Strategies

We evaluate four strategies for selecting which transformer layers to include in our feature extraction:

- **All layers**: Extract features from every layer (e.g., all 42 layers for Gemma-2-9B)
- **Every 2nd layer**: Sample at indices {2, 4, 6, …} reducing features by 50%
- **Every 5th layer**: Sample at indices {5, 10, 15, …} reducing features by 80%
- **Every 10th layer**: Sample at indices {10, 20, 30, …} reducing features by 90%

These strategies test whether dense sampling provides meaningful performance gains or if the model’s knowledge is sufficiently distributed that sparse sampling suffices. Sparser sampling dramatically reduces both memory requirements and inference time, making deployment more practical for resource-constrained environments.

##### Evaluation Protocol

For each sampling strategy, we train CNN-based probes using concatenated features from the selected layers. We measure three key metrics: accuracy, ECE, and wall-clock runtime for feature extraction and inference on the test set. Runtime measurements include both the forward pass through the transformer to extract activations and the probe inference time. All experiments use identical hardware (single NVIDIA H100 GPU) to ensure fair runtime comparisons.

We test on Gemma-2-9B-it (42 layers) and Llama-3.1-8B-Instruct (32 layers) as representative models with different architectures and layer counts. Each configuration is evaluated using 5-fold cross-validation to obtain confidence intervals.

#### 4.8.6 Refusal Experiments

We evaluate whether PING can recover task-relevant knowledge when surface-level refusals are induced either by prompting or by alignment fine-tuning. In all conditions, prompts are formatted with the model’s native chat template and the assistant turn is seeded with the fixed prefix “`Answer: ;`” to standardize the decoding surface.

##### Adversarial Prompting

We use a refusal-inducing instruction in the medical domain that explicitly directs the model to decline answering: “You are a general purpose assistant. However, you are not legally able to answer any medical-related question, even if it’s a multiple choice question. Instead, respond with, ‘Sorry, I cannot answer that. Please refer your question to a medical professional.’” This instruction is prepended to MedMCQA questions and options (A-D). Generative accuracy and refusal rate are computed under this prompt policy, with refusals marked incorrect.

##### Alignment Fine-tuning (DPO+LoRA)

To model refusals that are embedded in the weights rather than induced by a prompt, we fine-tune Gemma-2-9B-IT using Direct Preference Optimization (DPO; [30]) with LoRA adapters [31]. Each training pair consists of the same user question paired with (i) a safety-aligned refusal completion (*chosen*) and (ii) a contentful answer (*rejected* ). Both completions are prefixed with “`Answer:`” and formatted via the model’s chat template to ensure a stable surface form. The base model remains frozen; only adapters are trained. Hyperparameters follow standard small-adapter settings (e.g., LoRA rank *r*=16, dropout 0.1, and DPO scale *β*=0.1). We report refusal and accuracy metrics for this aligned model separately in the Results.

##### Refusal Parsing and Answer Extraction

For generated text, we declare a refusal if the output contains {any of sorry, cannot, can’t} (case-insensitive). Refused generations are counted as incorrect and no answer letter is extracted. For non-refusals, we extract the selected option using anchored patterns in priority order: `Answer: [A-D], Final answer: [A-D]`, or `The correct answer is [A-D]`; we additionally accept a markdown bold pattern (**A.) when present.

##### Probing Under Refusal

To test whether useful signal persists in internal representations despite refusals, we construct the exact inputs used for generation (chat-templated question with the assistant turn opened and seeded by “`Answer:` ) and extract activations for probing. We partition the evaluation corpus into three mutually exclusive splits derived from the generative run: **Correct** (non-refusal and correct), **Refused** (refusal), and **Incorrect** (non-refusal and incorrect). Unless otherwise noted, probes are trained on **Correct** and evaluated on **Refused**. Features are pooled at the start of the assistant turn (immediately after the “Answer:” anchor) and drawn from either a single best-performing layer or a small stack of late layers. Probes use a lightweight MLP with early stopping; metrics (accuracy and ECE) and stratified bootstrap confidence intervals follow Section 4.7.

##### Knowledge-Retention Subset

To isolate “knows vs. shows,” we further evaluate the probe on the subset of items where the generative model is known to be correct. We filter probe predictions by the baseline model predictions. Probe accuracy on this subset quantifies how often the internal signal still points to the correct option even when the aligned model refuses elsewhere.

### 4.9 Generative Baseline Methodology

To establish generative baselines for comparison with PING, we evaluate each model’s ability to directly generate correct answers through its standard decoding pathway. Our approach considers the constrained nature of multiple-choice questions to obtain both predictions and calibrated confidence scores.

#### Single-Token Generation

For multiple-choice questions with options A through D, we reformulate the task as predicting a single token. Each prompt is modified to end with “Letter of the correct answer:” to elicit a direct response. We then generate exactly one token using greedy decoding: *ŷ* = arg max_*v∈V*_ *p*(*v*| **x**), where *V* is the vocabulary and **x** is the input prompt. This approach avoids the complexity of parsing longer responses and provides a clean probability distribution over possible answers.

#### Token-to-Answer Mapping

The generated token *ŷ* is mapped to an answer choice {*A, B, C, D*} by first removing any prefix characters such as special tokens like or 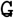, then converting to uppercase and matching against valid answer labels. Tokens that do not map to valid answer choices are treated as incorrect predictions with zero confidence, ensuring consistent handling of edge cases.

#### Generative Confidence Baseline

The model’s confidence in its prediction is taken as the softmax probability of the generated token: *p*_conf_ = exp(*z*_*ŷ*_)*/ Σ*_*v* ∈*V*_ exp(*z*_*v*_), where *z*_*v*_ are the logits for vocabulary token *v*. This provides a natural confidence score in [0, 1] that reflects the model’s uncertainty. For invalid tokens that do not map to answer choices A through D, we assign *p*_conf_ = 0 to indicate complete uncertainty, ensuring that our calibration metrics appropriately penalize the model for generating invalid outputs.

#### Temperature Scaling

We calibrate generative confidence scores using temperature scaling [32]. Given a vector of logits **z** and a scalar temperature *T >* 0, calibrated class probabilities are

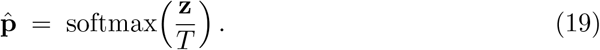

The temperature is chosen on a held-out calibration set by minimizing the negative log-likelihood (NLL):

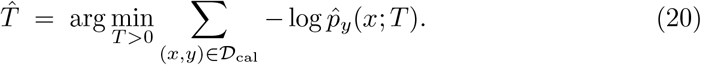

Temperature scaling is a one-parameter, post-hoc method that *preserves the predicted class* (the argmax), with *T >* 1 softening predictions (reducing overconfidence) and *T <* 1 sharpening them. In the binary case, given an uncalibrated probability *p* ∈ (0, 1) with logit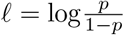and logistic sigmoid *σ*(*t*) = 1*/*(1+*e*^*−t*^), the calibrated probability is

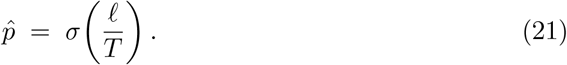

This parametric transformation preserves (for binary classification) the rank order induced by the logits while improving probability estimates.

#### Isotonic Regression

We also evaluate isotonic regression (IR) [33], a nonparametric calibration method that fits a monotone, piecewise-constant mapping *f* from model scores (e.g., probabilities) to calibrated probabilities. Given calibration scores {*s*_*i*_} and binary labels *y*_*i*_ ∈ {0, 1}, IR chooses a non-decreasing function *f* to solve

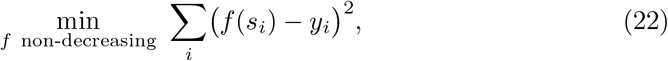

typically fit with the pooled-adjacent-violators (PAV) algorithm, yielding a stepwise (piecewise-constant) *f* . The learned mapping is applied to test scores as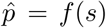, with outputs clipped to [0, 1] when used as probabilities. Unlike temperature scaling, IR can correct general (monotone) non-linear miscalibration patterns, at the cost of a higher risk of overfitting when the calibration set is small.

#### Evaluation Protocol

For each test example, the prompt is formatted according to the model’s instruction template and a single token is generated using the model’s standard inference pipeline. This token is then mapped to an answer choice or marked as invalid if it does not correspond to A, B, C, or D. Accuracy is computed as the fraction of correct predictions among all examples, while Expected Calibration Error is calculated using the extracted confidence scores. This approach ensures a fair comparison with PING by using the model’s native generation capabilities without modification, extracting confidence directly from the model’s output distribution, handling edge cases consistently, and avoiding potential confounds from longer generation such as explanation text that might affect answer extraction.

For models that refuse to answer, particularly in safety-critical domains like medicine, we record both the refusal rate and treat refused answers as incorrect with zero confidence. This allows us to quantify the impact of safety alignment on task performance while maintaining comparable metrics with PING. The single-token generation approach provides a principled baseline that captures both the model’s predictive accuracy and its calibration quality through its native decoding pathway.

## Data Availability

All data is available at https://github.com/tatonetti-lab/pingkit or publicly for MMLU

## 5 Appendix

### 5.1 Code and Tutorial Availability

The full PING framework implementation, including experiments, analysis scripts, and figure generation code, is available as an open-source Python package at https://github.com/tatonetti-lab/pingkit

The repository includes:

- **Installation instructions** for local and cloud environments.
- **Jupyter notebooks** demonstrating:
  – Extracting internal activations from HuggingFace-compatible LLMs.
  – Training probes on custom datasets.
  – Reproducing figures and tables from this manuscript.
  – Running layer sweeps and data efficiency experiments.
- **Tutorials** for integrating PING into existing NLP pipelines.
- **Pre-trained probe weights** for all models evaluated in this paper.

